# Mixed methods evaluation of using the ‘EMPathicO’ communication skills e-learning in primary care: “ *a super weapon to make the whole experience a bit better”*

**DOI:** 10.1101/2025.04.10.25325587

**Authors:** Emma Teasdale, Rachel Dewar-Haggart, Sebastien Pollet, Geraldine M Leydon, Hazel Everitt, Nadia Cross, Helen Atherton, Taeko Becque, Jennifer Bostock, Kirsty Garfield, Amy Herbert, Jeremy Howick, Paul Little, Christian Mallen, Leanne Morrison, Jacqui Nuttall, Matthew J Ridd, Michelle E Robinson, Beth Stuart, Jane Vennik, Nazrul Islam, Paul H Lee, Felicity L Bishop

**Affiliations:** Primary Care Research Centre, University of Southampton; School of Psychology, University of Southampton; PPIE; Population Health Sciences, University of Bristol; Centre for Academic Primary Care, University of Bristol; Leicester Medical School University of Leicester; Keele School of Medicine, Keele University; Southampton Clinical Trials Unit, University of Southampton; Wolfson Institute of Population Health, QMUL

**Author notes:** Corresponding author: Dr Emma Teasdale University of Southampton Aldermoor Health Centre Aldermoor Close Southampton SO16 5ST.

**Keywords:** Primary Health Care, Empathy, Optimism, Health Communication, Digital Technology, nested qualitative study

## Abstract

**Objectives:** To examine primary care practitioners’ experiences and use of EMPathicO e-learning to enhance communication of clinical empathy and realistic optimism.

**Design:** Mixed methods evaluation incorporating thematic analysis of qualitative interviews and quantitative analysis of EMPathicO usage patterns and practitioner survey data.

**Setting:** Cluster randomised controlled trial of EMPathicO in general practices in England and Wales.

**Participants:** Primary care practitioners allocated to the intervention arm.

**Analysis:** Thematic analysis of qualitative data explored experiences of undertaking EMPathicO and implementing change in subsequent consultations. Descriptive quantitative analysis of EMPathicO usage and practitioner-reported survey data examined practitioner engagement with the e-learning. These parallel analyses were integrated using a triangulation protocol to explore convergence, complementarity and dissonance between the datasets.

**Results:** 23 interviews (16 initial and 7 follow up) across 14 GP practices were undertaken with 11 GPs, 1 nurse practitioner, 3 physiotherapists and 1 physician associate, purposively sampled for diversity from the 115 participants randomised to receive EMPathicO in the trial. Interviewees were positive about EMPathicO, perceiving it as convenient and manageable (approx. 75 minutes online), informative, important, and relevant to their consultations. Over 95% of the 115 EMPathicO practitioners completed the e-learning modules, set goals and felt motivated to adopt EMPathicO communication skills following the e-learning. Interviewees appreciated the autonomy of setting personal goals; found their chosen empathy and optimism goals feasible to incorporate into everyday practice without lengthening consultations and felt such changes led to more positive interactions that were mutually beneficial for practitioners and patients. Aside from their own personal benefit some felt EMPathicO would be particularly helpful if integrated into existing training programmes. One interviewee described how they did not adopt the EMPathicO communication skills (despite feeling positive towards the e-learning overall) due to nearing retirement and another described not using specific tools within the e-learning (e.g. goal setting) because they did not fit with their preferred learning style. Additional content on communicating clinical empathy and realistic optimism flexibly in some situations (e.g., remote consultations especially telephone due to limitations on non-verbal communication) would be welcomed.

**Conclusions:** Practitioners across the multidisciplinary primary care team found completing EMPathicO to be a positive experience, manageable in the current pressurised clinical context and worthwhile, perceiving it to enhance their communication skills. They felt it benefitted both them and their patients and could also be particularly helpful within GP training and medical education settings. These important findings would have been missed if the mixed methods evaluation had not been incorporated into the trial. If widely disseminated, EMPathicO is likely to be well-received by primary care practitioners and straightforward to integrate into everyday practice.

**Summary:** *What is already known:*

- Effective practitioner-patient communication, including clinical empathy can help enhance healthcare interactions, patient outcomes and self-management and improve practitioner job satisfaction and reduce burnout.
- Existing clinical empathy training can be too lengthy for busy primary care clinicians, is commonly delivered face to face limiting accessibility, and has not usually been offered across the multidisciplinary team.
- EMPathicO is rigorously developed, evidence-based brief communication skills e-learning to enhance communication of clinical empathy and realistic optimism.

*What this study adds:*

- Primary care practitioners across the multidisciplinary team valued EMPathicO, finding it relevant, accessible, manageable in the context of busy clinical practice and a positive worthwhile experience.
- Practitioners felt motivated to adopt EMPathicO communication skills and most reported doing so, perceiving benefits for themselves and their interactions with patients, without lengthening consultation length.
- If widely disseminated, EMPathicO is likely to be very well-received by primary care practitioners and straightforward to integrate into everyday practice.

## INTRODUCTION

Effective practitioner-patient communication can help to enhance healthcare interactions, improve self-management and patient outcomes. ^1–3^ Showing clinical empathy (where the practitioner seeks to understand and respond to the patient’s perspective) and realistic optimism (conveying a positive message where appropriate) are essential components of effective healthcare communication ^4^ ^5^ that can benefit patients in terms of satisfaction with care ^6^ ^7^ and practitioners in terms of job satisfaction and reduced burnout. ^8^ ^9^ Previous quantitative and qualitative systematic reviews of empathy-focused training have suggested that such training can improve practitioner empathy ^10^, professional development, and patient care. ^11^ However current training is time-consuming for busy primary care practitioners (e.g. over 10 hours); typically delivered face to face; not offered across the multidisciplinary primary care workforce and does not incorporate realistic optimism. ^12^

EMPathicO is a rigorously developed, evidence-based, theoretically-grounded communication skills e-learning for primary care practitioners to enhance their communication of clinical empathy and realistic optimism within consultations. ^12–18^ It comprises three brief interactive modules covering clinical empathy, realistic optimism and osteoarthritis (included as an example of communicating empathy and optimism in a particular context). These modules are followed by reflection and goal setting activities, in which practitioners consider what works well/less well in their consultations; select 2-3 empathy and optimism goals from 15 suggested goals or create their own; plan how and when to implement selected goals in subsequent consultations (Figure 1). Each module incorporates multiple behaviour change techniques and can be completed separately or all together. For full description as per TIDieR see supplementary materials. The Talking in Primary care (TIP) cluster-randomized controlled trial assessed the effectiveness and cost-effectiveness of EMPathicO on patients’ musculoskeletal (MSK) pain and enablement. It was undertaken in primary care in England and Wales and involved 233 practitioners and 1,682 patients from a diverse range of 53 GP surgeries randomised to undertake EMPathicO (intervention) or consult patients as usual (control).

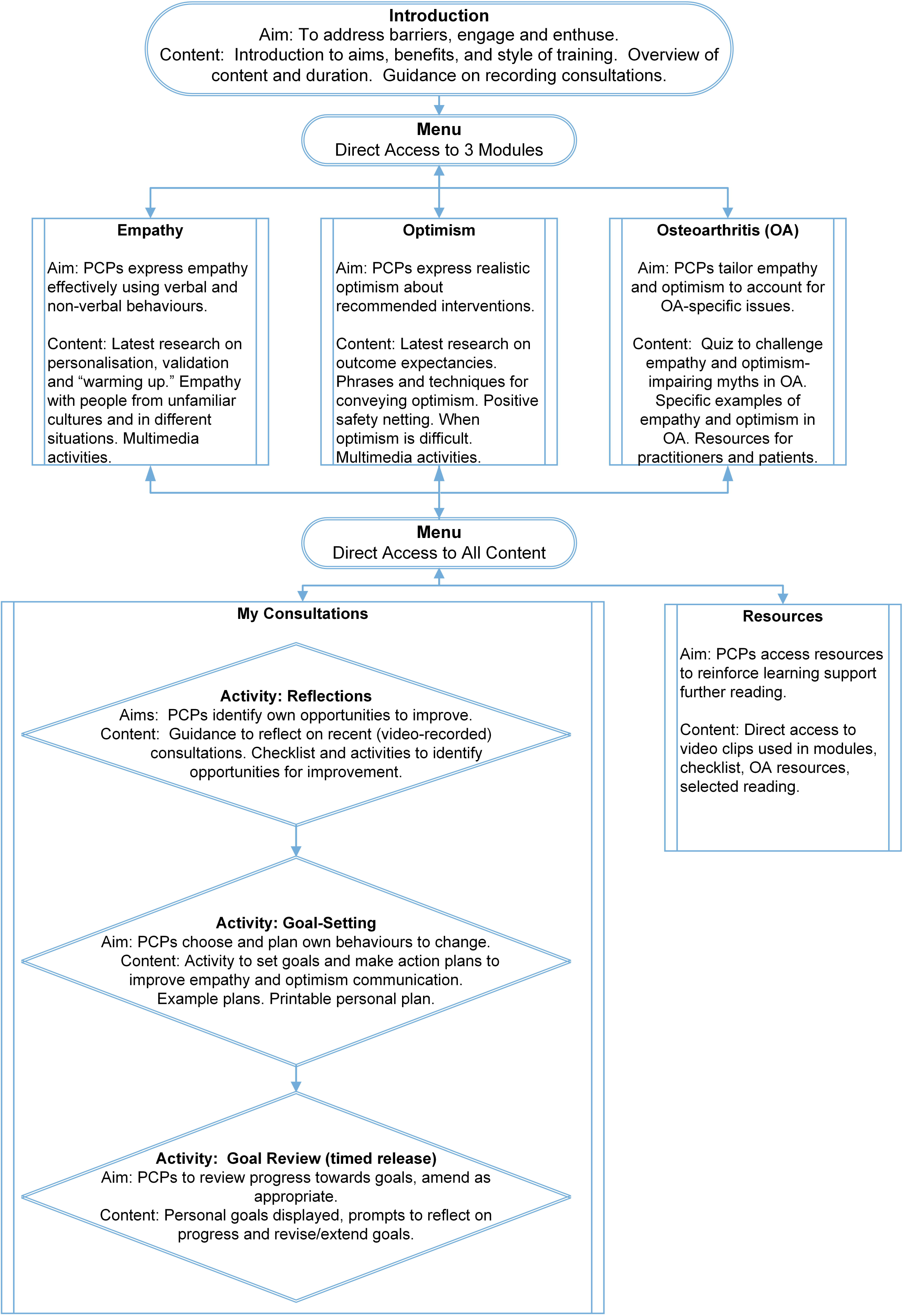
Figure 1

Conducting mixed methods evaluations alongside trials can add depth by exploring trial participants’ views and experiences of an intervention and its implementation and identifying contextual factors that may help explain observed trends in trial outcomes. ^21^ Findings from the TIP trial are reported in full elsewhere and suggest brief e-learning for primary care practitioners in clinical empathy and realistic optimism is safe, probably cost-effective, and significantly enhances practitioner self-efficacy over a sustained period but found no effect on patient outcomes. The aim of this paper is to examine practitioners’ experiences and use of EMPathicO and understand how and why using EMPathicO may benefit practitioners. This analysis was conducted before the trial results were available.

## METHODS

### Design

A (QUAL + quan) mixed methods approach ^22^ was used, combining a core qualitative component (thematic analysis of qualitative interviews) with a supplementary quantitative component (analysis of EMPathicO-usage data and survey data on practitioner beliefs about, intentions towards, and implementation of EMPathicO skills). Each component proceeded in parallel; insights from the qualitative and quantitative data analyses were systematically compared following the triangulation protocol ^23^ to offer a comprehensive analysis of practitioners’ views, experiences, and use of EMPathicO. Qualitative interviews were also conducted to examine the views and experiences of control arm practitioners and practice staff about participating in the TIP trial (to be reported elsewhere).

The qualitative component is reported in accordance with O’Brien’s standards for reporting qualitative research. ^24^ Qualitative team members comprised two female qualitative research fellows (EJT, RDH), a female health psychologist (FLB), a female general practitioner (HAE), a male research fellow (SP) and a female medical sociologist (GL) who met weekly to discuss data collection and collaborate on data analysis. Ethics approval was obtained from the South Central - Hampshire B Research Ethics Committee (ref: 22/SC/0145).

### Participants and Recruitment

#### Practitioners

115 of the 233 practitioners participating in TIP were randomised to the intervention arm and emailed login credentials to access EMPathicO. ^20^

#### Interviewees

As general practices completed trial patient recruitment, the qualitative team emailed a qualitative study invitation (cover letter, information sheet and consent form) to a purposive sample of practitioners chosen to incorporate variety in profession, gender, age, ethnicity, geographical location and years of experience. By sampling for variety, we helped ensure that the qualitative findings captured the range of participants’ experiences. All practitioners who expressed interest and completed the consent form were interviewed via MS Teams at a mutually convenient time. Approx. 3-6 months after their initial interview, practitioners were invited to a follow up interview. EJT conducted 23 interviews (16 initial and 7 follow up) across 14 GP practices between March 2023 and June 2024 (Table 1).

**Table 1:**
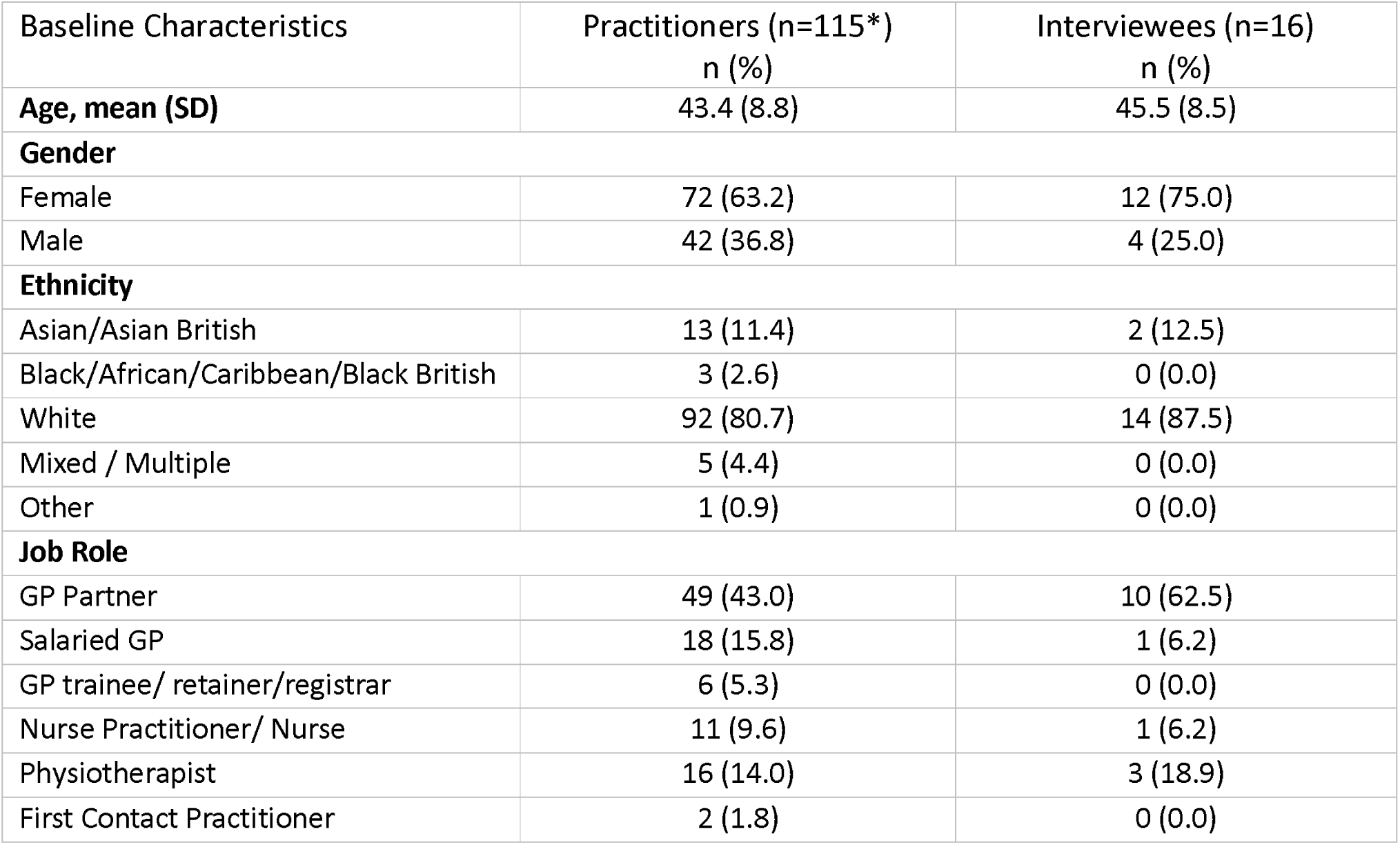
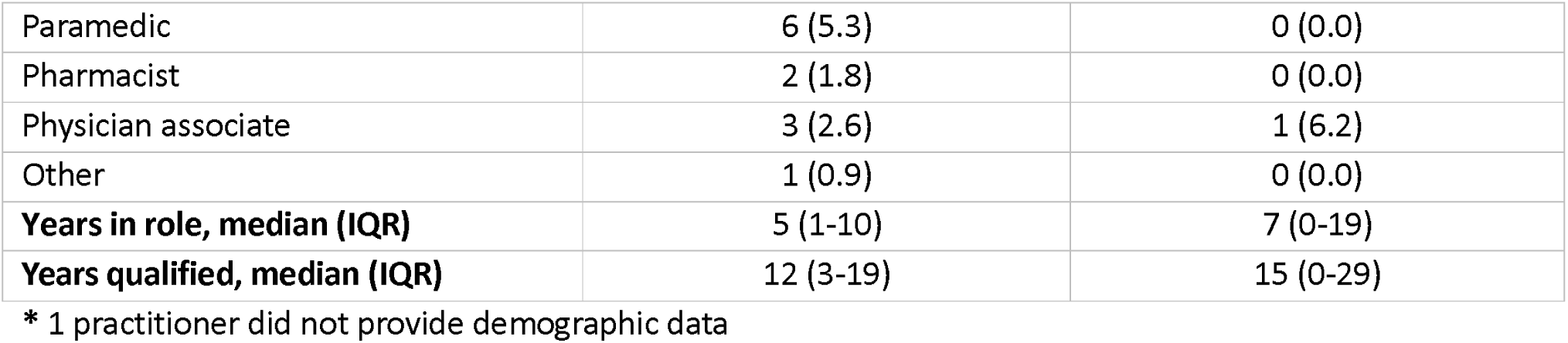
Practitioner and interviewee characteristics

| Baseline Characteristics | Practitioners (n=115*)<br>n (%) | Interviewees (n=16)<br>n (%) |
| --- | --- | --- |
| **Age, mean (SD)** | 43.4 (8.8) | 45.5 (8.5) |
| **Gender** |  |  |
| Female | 72 (63.2) | 12 (75.0) |
| Male | 42 (36.8) | 4 (25.0) |
| **Ethnicity** |  |  |
| Asian/Asian British | 13 (11.4) | 2 (12.5) |
| Black/African/Caribbean/Black British | 3 (2.6) | 0 (0.0) |
| White | 92 (80.7) | 14 (87.5) |
| Mixed / Multiple | 5 (4.4) | 0 (0.0) |
| Other | 1 (0.9) | 0 (0.0) |
| **Job Role** |  |  |
| GP Partner | 49 (43.0) | 10 (62.5) |
| Salaried GP | 18 (15.8) | 1 (6.2) |
| GP trainee/ retainer/registrar | 6 (5.3) | 0 (0.0) |
| Nurse Practitioner/ Nurse | 11 (9.6) | 1 (6.2) |
| Physiotherapist | 16 (14.0) | 3 (18.9) |
| First Contact Practitioner | 2 (1.8) | 0 (0.0) |
| Paramedic | 6 (5.3) | 0 (0.0) |
| Pharmacist | 2 (1.8) | 0 (0.0) |
| Physician associate | 3 (2.6) | 1 (6.2) |
| Other | 1 (0.9) | 0 (0.0) |
| **Years in role, median (IQR)** | 5 (1-10) | 7 (0-19) |
| **Years qualified, median (IQR)** | 12 (3-19) | 15 (0-29) |

\* 1 practitioner did not provide demographic data

### Qualitative Data Collection

Written (electronic) consent was elicited prior to interview. The interviewer used a topic guide (supplementary materials) comprising open-ended questions designed by the qualitative team and PPIE collaborators to explore interviewees’ experiences and perspectives in-depth and to draw out novel and unforeseen insights. The topic guide was informed by key domains from Normalisation Process Theory (NPT) ^25^, which provides a framework for identifying factors and processes likely to hinder or enable widespread implementation of new practices into routine practice outside of study processes. Questions tapped the four constructs of NPT: coherence (people making sense of the processes), cognitive participation (engaging with the process), collective action (work that is required to operationalise the process), and reflexive monitoring (reflecting on and appraising new working practices).

Interviews were audio-recorded (using MS Teams). Field notes were taken to capture the interviewer’s reflections, and any aspects not captured by the recording. Mean interview duration was 36 minutes (range 27 to 48). On completing each interview, interviewees were thanked, debriefed and offered a copy of their transcript and study findings, when available. Interviewees were reimbursed for their time as part of the practice research costs payment. Data collection stopped on reaching thematic saturation (i.e., themes were rich, well-developed, and understood) and a thorough, credible analysis in relation to our aims had been achieved. ^26^

### Quantitative Data Collection

Quantitative usage data (e.g., number and duration of logins, pages visited) were automatically recorded by the LifeGuide+ platform ^27^, used to create and host the EMPathicO e-learning. Practitioners also completed online surveys via Qualtrics (Qualtrics, Provo, UT) assessing their beliefs about, intentions towards, and self-reported change in communicating empathy and optimism in consultations.

### Data analysis

#### Interviews

All interviews were professionally transcribed verbatim and identifiable information removed. Data collection and initial analyses proceeded iteratively (i.e., coding started after the first few interviews and informed subsequent interviews). Thematic analysis ^28^ was used to explore the data. One author (EJT) read and reread the transcripts to familiarise herself with the data. Codes were derived inductively from the data and grouped together to produce an initial coding frame. To enhance the quality and credibility, a more detailed coding frame was then created to ensure transparent and systematic coding. Codes and theme/sub-theme definitions were discussed with, and iteratively developed by, the qualitative team and PPIE contributors to offer diverse inferences and to avoid idiosyncratic interpretations. A negative case analysis was carried out to capture anomalous cases and enable us to identify important, but infrequent, views. Although analysis was primarily inductive (i.e., data driven), normalisation process theory ^25^ was consulted during analysis to aid interpretation. NVivo (NVivo version 14, Lumivero, 2023) was used to manage data, implement, and record coding.

#### Usage and survey data

A descriptive quantitative analysis examined the extent to which practitioners engaged with EMPathicO during the TIP trial in terms of the number of times practitioners logged in; total time spent logged in; total time spent logged into the core modules; and proportion of PCPs completing core modules, reflecting on consultations and setting goals (analysed by SP). Survey data on self-reported change in communicating empathy and optimism in consultations were analysed descriptively by TB.

#### Data integration

Key themes in the qualitative data and trends in the quantitative usage and survey data were mapped in matrices and diagrams (Table 2). Adopting a triangulation protocol ^23^, these datasets were integrated and examined in terms of the extent to which they agreed (convergence), complemented one another (complementarity) or contradicted each other (dissonance), allowing a comprehensive understanding of practitioners’ experiences of EMPathicO to be developed.

**Table 2:** Triangulation matrix mapping qualitative themes onto key quantitative findings

| Qualitative themes | Quantitative (usage) findings |  |  |  |  |
| --- | --- | --- | --- | --- | --- |
|  |  | High rates of training completion | Brief overall training duration | High rates of variable goal setting |  |
|  |  | Most practitioners (over 95%) completed the training. | Most practitioners took between 30 minutes and 120 minutes to complete it and did this overall more than one session. | Most practitioners (over 95%) fully set 2-3 goals and across the group they set a variety of different empathy and optimism goals. Most practitioners reported making changes to subsequent consultations. |  |
|  | **EMPathicO was practically, intellectually, and clinically valuable** | Interviewees found completing EMPathicO *informative and relevant* to primary care consultations.<br>*Interviewees* thought showing empathy and optimism was important (*significant*) for effective practitioner-patient communication. | Interviewees found EMPathicO *manageable* to complete. Convenience of the online format, succinct content and modular structure were important aspects in making EMPathicO feel feasible to do. |  | Interviewees thought EMPathicO would be helpful for less experienced practitioners and students/trainees |
|  | *Feeling motivated to adopt EMPathicO communication skills following e-learning* |  |  | Interviewees appreciated being able to set personal goals, found their chosen empathy and optimism goals feasible to incorporate into everyday practice without lengthening the consultation and felt making such changes incurred benefits for both patients and practitioners. |  |
|  | *Implementing EMPathicO communication skills flexibly to suit diverse primary care contexts* |  |  |  | Interviewees felt that showing empathy and optimism was more challenging in some circumstances. |

### Data and Code Sharing

Data and code are available on request. Requests for deidentified participant data may be submitted to the University of Southampton data repository, quoting doi: [to be added on acceptance]. Requests would be subject to review by a subgroup of the trial team. Access to anonymised data may be granted following this review, subject to conditions including ethical approval, qualifications, and aims consistent with the original purpose of the study. All data-sharing activities would require a data-sharing agreement. Code will be shared via GitHub repository.

### Patient and Public Involvement

A diverse group of 6 patient and public collaborators (PPIE) were involved in this research. They had bi-monthly meetings over the course of the trial, contributing to pilot qualitative interviews, qualitative analysis and interpretation of findings (see Bishop et al, (2025) for further details).

## RESULTS

Our analyses produced 3 main themes (Figure 2) related to practitioners’ views on and experiences of undertaking and implementing EMPathicO:

1. ***EMPathicO was practically, intellectually, and clinically valuable***.
2. ***Feeling motivated to adopt EMPathicO communication skills following e-learning***.
3. ***Implementing EMPathicO communication skills flexibly to suit diverse primary care contexts***.

These themes and ***subthemes*** are explored below by presenting selected illustrative, anonymised quotations and incorporating findings from the usage and survey data analysis.

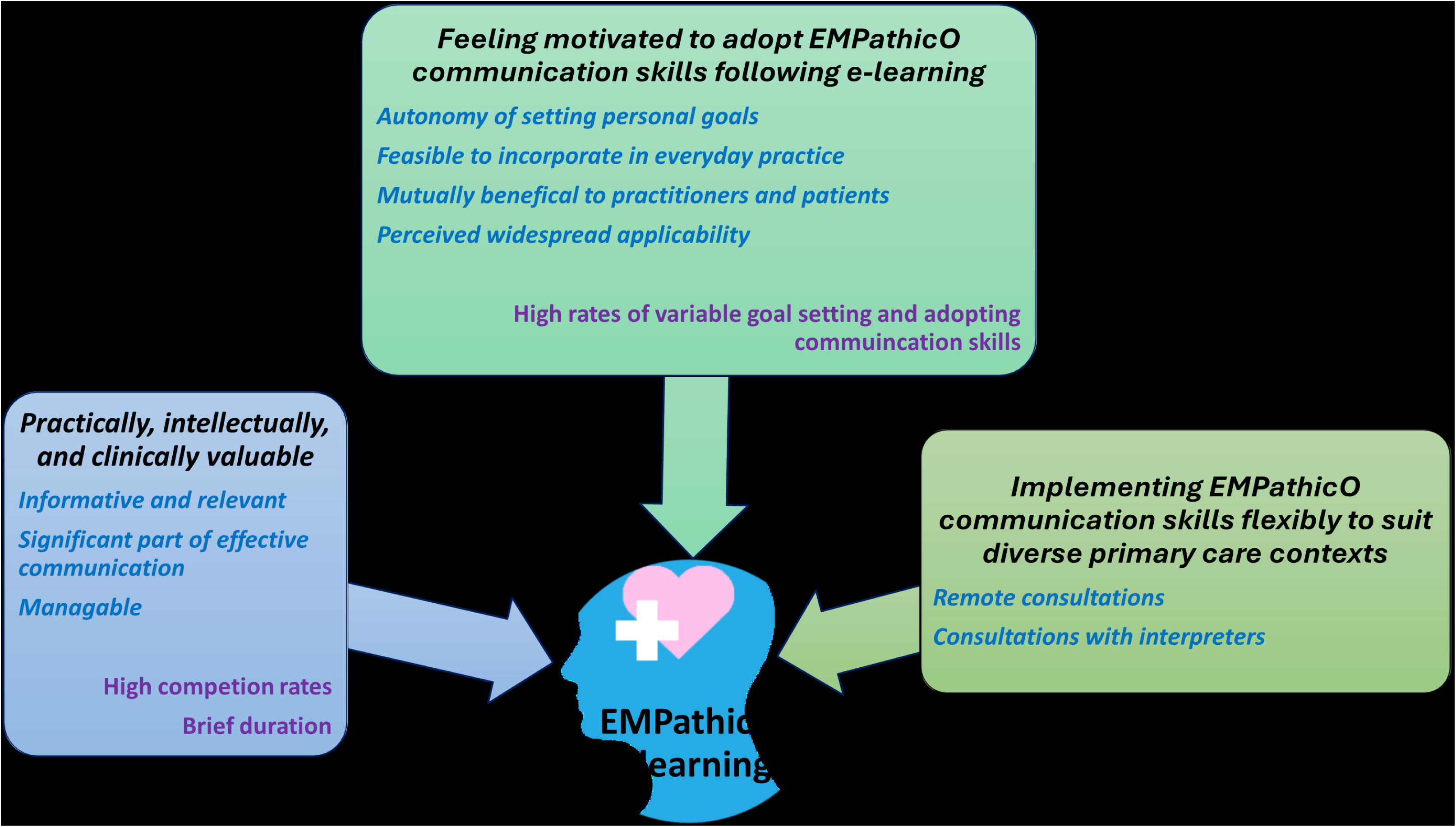
Figure 2

### EMPathicO was practically, intellectually, and clinically valuable

The predominant view was that undertaking and implementing EMPathicO was positive and worthwhile. Key messages were well received. Interviewees described EMPathicO as clear, ***informative and relevant to primary care***. They particularly liked the practical tips and advice and found the visual aspects of EMPathicO useful, such as “*watching the [simulated] videos with the patient of how they did it, how the communication went.” (P1, Female Physio)*. They also found the inclusion of the evidence-base for taught techniques compelling, indicating *“it wasn’t just somebody’s idea, there was some more information about why doing certain things might be a good idea” (P31, Female GP)*.

> *“Simple, clear, just the right length, practical, and I like the video clips, because that emphasised what the training’s about….I think there was a bit where the GP does what he normally does and then just changes how he would do it after, so how he used some of the phrases and how he also echoed some of the patients’ concerns verbally…. the fact I remember it after this length of time is saying something but It’s talking about something that’s relevant to what I do day to day.” (P27, Female GP)*

Interviewees also reflected on the importance of EMPathicO which seemed to relate to the dominant view that ***showing clinical empathy and realistic optimism is a significant part of effective communication*** in day-to-day primary care.

> *“That’s a very important part of our consultation. If we’re not going to show positivity to the patient, how can we ask them to stay positive? It has to come from us to the patients, and also empathy. These two are really important to have a successful outcome and support the patient.” (P1, Female Physio)*
>
> *“Empathy is absolutely paramount to what we do. If you don’t have empathy, you’re burnt out, and you shouldn’t be doing the job. Positivity, yes, it’s very incredibly helpful and I think, well, you’d have to look into longitudinal stuff, but it almost certainly would lead to more positive outcomes than negative outcomes.” (P32 Male GP)*

The perceived relevance and importance of EMPathicO was also reflected in the usage data, which showed high practitioner engagement during the trial. Of the 115 practitioners allocated to the intervention arm, the majority (n=111, 96.5%) logged into and accessed the e-learning. All 111 practitioners who logged into EMPathicO completed all three modules and 96.4% (n=107) completed the reflections and goal setting activities (Table 3).

**Table 3:** Practitioner EMPathicO completion rates

| EMPathicO completion rates | Practitioners (n=111)<br>n (%) |
| --- | --- |
| Clinical empathy module | 111 (100) |
| Realistic optimism module | 111 (100) |
| Osteoarthritis module | 111 (100) |
| Reflection and goal setting activities | 107 (96.4) |

Interviewees across the multidisciplinary primary care team found doing the e-learning ***manageable***, because it was succinct with a convenient, easy to use online format and modular structure. Interviewees appreciated having the flexibility to access EMPathicO “in your own time and in your own space” (P14, Male GP) and to “work through it at your own pace” (P8, Female Physician Associate), as well as having it as a resource to revisit as required. The overall duration of the e-learning was considered appropriate to fit into current workloads; “it wasn’t anything too onerous” (P12, Female Nurse). They found the modules easy to navigate and liked that it was “in quite good bitesize chunks that you didn’t feel that you were wading through a lot of information” (P5, Female GP) that allowed them to “dip in and out” (P25, Female GP) or complete all at once.

The manageability of EMPathicO was also evident in the quantitative data on duration and patterns of use. Most practitioners (73.0%, n=81) logged into EMPathicO more than once; median was 2 logins, range 1-11. Median overall time (across all logins) spent completing the 3 modules and activities was 49 minutes. Median overall time spent logged into EMPathicO (across all logins, including time completing questionnaires) was 75 minutes, with most practitioners (n=85, 76.6%) spending between 30 and 120 minutes logged into EMPathicO.

### Feeling motivated to adopt EMPathicO communication skills following e-learning

Interviewees described feeling inspired to make changes to their communication within consultations following EMPathicO and this in part related to a sense of agency from having the ***autonomy to set personal goals.*** They felt that the non-prescriptive approach to goal setting in EMPathicO helped to render it feasible to both set and implement their chosen goals in everyday practice as they could focus on personally relevant skills.

> *“I like achieving my own goals I set for myself. I think, if we leave that bit out from the training, it becomes less - maybe you adhere to the whole training and continue with it, but if you set your own goals, it becomes more individual for you to continue with it. I think that’s an important part.” (P1, Female Physio)*

Quantitative data indicated that practitioners typically felt motivated to adopt EMPathicO communication skills. Most practitioners (n=107, 96.4%) reflected on at least one prior consultation (out of suggested maximum of 5 consultations) and made complete goal-setting plans including an intended start time (n=106, 95.5%). The perceived applicability of the empathy and optimism goals and the importance of being able to set personal goals was further reflected in the variety of goals chosen by practitioners (Table 4). Practitioners selected a range of goals from across the 15 suggested in EMPathicO, with only one practitioner creating their own goal.

**Table 4:**
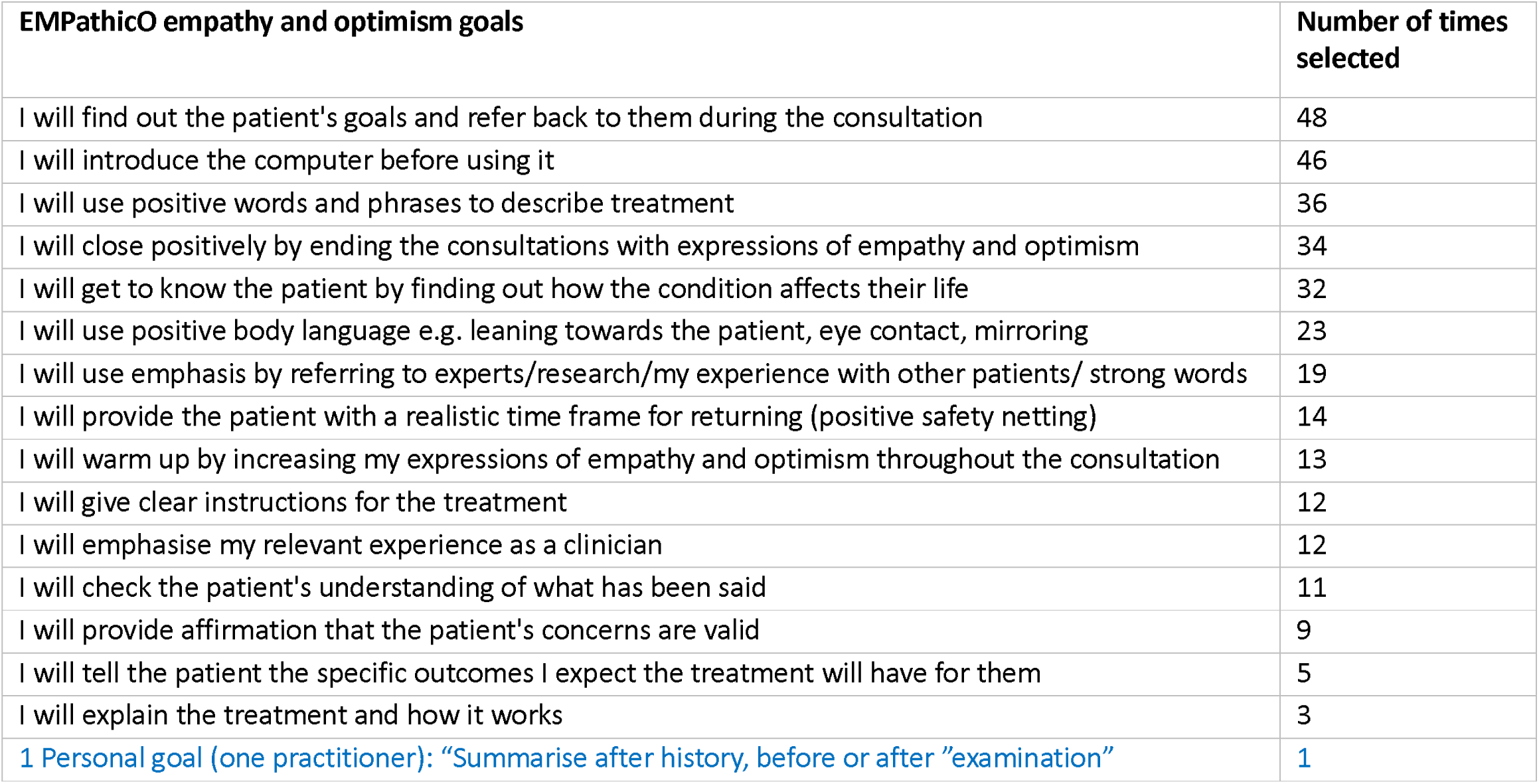
Frequency/variability of selected goals across intervention arm practitioners

Interviewees also described setting a range of different empathy and optimism goals and explained how these goals resonated with them due to perceived novelty or potential enhancement of existing skills. Providing a range of goals for practitioners to choose from worked well and allowed consideration of the diverse ways in which empathy and optimism can be construed and communicated.

> *“That was a novel approach, to me, as well as the rephrasing, the ending of the consultations, that, ’Try this, but if it doesn’t get better, come back.’ Again, you can just hear yourself saying it all the time, and actually, when you think about it - and the training made me think about it - you do think, it is a very negative way of ending a consultation, and the person will go away thinking, okay, so if it doesn’t work, I’ll just go back and see them, whereas if you go away thinking, it sounds like a good treatment; I know that I can go back, but actually, I’m hoping that, over the next few weeks, this will just get better, it’s a very different way of leaving the consultation. Those were quite novel things for me.” (P14, Male GP)*

One practitioner selected goals but did not create a plan or start time. Four practitioners did not select any goals. One interviewee described how they skipped the goal setting activities because “It’s just not me.” (P6, Female GP) and suggested that such activities did not fit with their preferred learning style.

> *“It depends on your character, how you do stuff, doesn’t it? I’m not very driven by all these goals, and nonsense like that. I’m old. My generation grew up with, ’This is what it is. Get on with it!’ (P6, Female GP)*

Interviewees felt their chosen empathy and optimism goals were ***feasible to incorporate into everyday practice without lengthening the consultation***. A dominant view was that making these changes was not *“trying to reinvent the wheel” (P5, Female GP)* or implement a new consultation style but rather offered a subtle evidence-based way to think about nuance of communication *“it’s finding little changes, my gold nuggets” (P4, Female GP)* , thereby enhancing existing consultation styles. Consequently, interviewees felt that their chosen goals were easy to embed in practice and minimally impacted their workload. This experience persisted and was described during follow up interviews up to 14 months after initial use of EMPathicO.

> *“I don’t feel it increased the workloads. I think because being empathetic is part of my role anyway, I think that was something I would do, it’s just using different ways of communicating with a patient, but it didn’t add to any extra time with the patients. (P3-Female Physio)*
>
> *“It’s become part of my normal consultations now. So, it’s ingrained.” (P14, Male GP)*

Making EMPathicO changes was also viewed by practitioners as ***mutually beneficial*** for them and their patients. Interviewees felt that implementing their empathy and optimism goals in consultations was “*a kind of super weapon to make the whole experience a bit better” (P13, Female GP)*. They commonly described how making changes had led to positive patient interactions which made them feel better post consultation e.g*., “the positiveness of the approach makes me feel positive” (P31, Female GP).* Interviewees also suggested how making changes following EMPathicO had helped them to reach mutually agreeable management plans more readily and aided them in managing challenging consultations.

> *“I found them very helpful. As I’ve alluded to, I’ve noticed an improvement in my consultation flow, and my relationship with the patients because of them. I find we reach a mutually acceptable management plan sooner, and I’ve noticed that it is - yes, patient outcomes tend to be better because they are willing to try the non-pharmacological, non-surgical therapies, and put a bit more time into them than they might have done previously.” (P32, Male GP)*

Quantitative data indicated that practitioners typically felt motivated to implement their selected empathy and optimism goals into everyday clinical practice following EMPathicO. At 8 weeks post-randomisation, 76.7% of practitioners (56/73 questionnaire respondents) reported making changes to their routine practice since completing EMPathicO. Similarly, at 34 weeks post-randomisation, 65.1% (54/83 questionnaire respondents) reported making changes to their routine practice since completing EMPathicO.

Interviewees’ appreciation of EMPathicO was further evident in its ***perceived widespread applicability***. EMPathicO appealed to a range of different interviewees from across the multidisciplinary primary care workforce, including very experienced practitioners who felt that EMPathicO was a valuable approach for refreshing existing communication skills “*as you get a bit longer in the tooth in the profession it’s revitalising to go through this kind of training” (P27, Female GP).* EMPathicO was also commonly viewed as a valuable learning resource for less experienced practitioners and for undergraduate and/or postgraduate medical and healthcare educational programmes, providing “*a good opportunity to get people whilst they’re learning early on in their career.” (P14, Male GP).* Interviewees also reflected on how their empathy and optimism goals were applicable in a wide variety of consultations (beyond patients with MSK).

> *“You have to judge it on your patient that you’re with. Definitely for people with pain and MSK, I suppose it makes the most sense, because often there isn’t a tablet that’s going to necessarily make it all better… but even if someone’s got a chest infection, or a cough, or the flu, or anything, you can try and encourage them to be more positive about it. I mean, even if someone’s got cancer and they’re dying, actually you can still use the empathetic-ness and the positive bits and finding the silver lining bits. (P4, Female GP)*

### Implementing EMPathicO communication skills flexibly to suit diverse primary care contexts

While EMPathicO was experienced as relevant and helpful for patients with diverse health needs, interviewees also highlighted circumstances where they would appreciate additional support within EMPathicO. Although EMPathicO does contain information/advice on showing empathy and optimism in remote consultations, interviewees felt that their ability to show empathy on the telephone was sometimes limited as “you can’t use all the non-verbal communication skills” (P1, Female Physio), or due to technical issues/poor connection and that some remote consultations can be less discursive, more task-based.

> *“I find over the telephone is much more of a fact-finding mission in terms of like your history-taking or whatever. When you see a patient face-to-face, you do tend to digress more and have more of a kind of dialogue and interaction.” (P12, Female Nurse).*

Interviewees also reflected on the challenges of communicating empathy in consultations when using an interpretation service, in particular expressing doubts about the ability of the interpreter to convey the practitioners’ empathy adequately.

> *“You also don’t know if the actual interpreter is actually interpreting your trying to be empathetic. So I do think they often don’t because I can sometimes understand bits of it, and it’s not being interpreted word for word.” (P25,Female GP )*

Similarly, interviewees described how showing realistic optimism still felt difficult in some consultations where there might be lots of upset and anger because “*there might not always be an optimistic outlook with some things” (P3, Female Physio)* .

## DISCUSSION

Interviewees were positive about the EMPathicO e-learning, finding it convenient, manageable, informative and relevant to their consultations. The focus of EMPathicO (conveying empathy and optimism to patients) was seen as vital for effective practitioner-patient communication. Interviewees felt motivated to adopt EMPathicO communication skills following the e-learning: they appreciated the autonomy of setting personal goals; found their chosen empathy and optimism goals feasible to incorporate into everyday practice without lengthening consultations and felt such changes led to more positive interactions which were mutually beneficial for practitioners and patients. These qualitative findings were also reflected in the quantitative data that showed over 95% of the 115 EMPathicO practitioners completed the e-learning modules, set goals, felt motivated to adopt EMPathicO communication skills following the e-learning and reported that using EMPathicO had been practically useful in enhancing their communication skills in subsequent consultations. Interviewees also felt EMPathicO had broad appeal and applicability outside of the trial setting and could be especially helpful if integrated into existing training programmes. One interviewee described how they did not adopt the EMPathicO communication skills (despite feeling positive towards the e-learning overall) due to nearing retirement and another described not using specific tools within the e-learning (e.g. goal setting) because they did not fit with their preferred learning style. Additional content on communicating clinical empathy and realistic optimism flexibly in some situations (e.g., remote consultations especially telephone due to limitations on non-verbal communication) would be welcomed.

A key strength of this evaluation was our mixed methods approach. By analysing qualitative interviews drawing on normalisation process theory and quantitative usage and survey data, the rates of engagement with EMPathicO along with practitioners’ in-depth experiences could be examined. This work provides new insights into primary care practitioners’ views and experiences of undertaking and implementing communication skills e-learning in everyday clinical practice. A diverse sample of practitioners were included in the qualitative study, and all EMPathicO practitioners were included in the usage analyses, ensuring that our findings are not based on a narrow subset of participants. A further strength was the multidisciplinary nature of the qualitative team including input from patient collaborators that meant that we approached the data from diverse perspectives, achieving richer insights than would otherwise have been possible. It is also worth noting that practitioners from across the primary care workforce talked positively about EMPathicO in interviews conducted at least 3 months after initial use (first interviews were 3-to 10-months following EMPathicO and follow-up interviews were up to 14 months following EMPathicO). Practitioners who were highly engaged and positive about EMPathicO may have been more likely to participate in qualitative interviews. However, given how well all practitioners engaged in EMPathicO this is unlikely to account for the qualitative findings. It is also possible that practitioners in the trial may be more interested in communication than non-trial participants.

Our findings map to key concepts from Normalisation Process Theory. ^25^ EMPathicO was ‘coherent’ to practitioners, in that communicating empathy and optimism in consultations made sense. Practitioners demonstrated ‘cognitive participation’ in EMPathicO, in that they understood the importance of effective practitioner-patient communication and its potential benefits, appreciated its ease of use and therefore committed to it. To facilitate ‘collective action’, practitioners reflected on the wide applicability of EMPathicO across the primary care workforce and beyond. Our findings are also consistent with evidence from a systematic review of experiences of empathy training in healthcare, which found that empathy training was beneficial to its participants, and they gained a deeper recognition of the positive impact of empathic care. ^11^

This analysis helps us to understand why the TIP trial found a sustained increase in practitioner confidence to communicate clinical empathy and realistic optimism to their patients. By providing clear, brief, evidence-based information in multiple formats and supporting autonomous behaviour change, EMPathicO empowered practitioners to make specific goal-driven changes to their communication behaviours. Beyond aiding interpretation of the trial results, this mixed methods evaluation has generated recommendations to support wider discussions with diverse stakeholders on enabling wider use of EMPathicO. For example, inclusion of EMPathicO in medical undergraduate and postgraduate educational programmes and as CPD for experienced practitioners. The perceived suitability of EMPathicO for wider use is supported by the concept of the habit formation at an early stage being preferable to trying to change ingrained but unhelpful habits among later-stage professionals. Future work should develop resources to implement recommended actions to support wider use of EMPathicO and to consider providing additional support around conveying empathy and optimism in remote consultations. If rolled-out, EMPathicO is likely to be well-received by primary care practitioners and straightforward to integrate into everyday practice.

## Additional Information

### Ethics Approval

Ethics approval was obtained from the South Central - Hampshire B Research Ethics Committee (ref: 22/SC/0145) and all participants gave informed consent before taking part.

### Transparency

The guarantor (F L Bishop) affirms that the manuscript is an honest, accurate, and transparent account of the study being reported; that no important aspects of the study have been omitted; and that any discrepancies from the study as originally planned and registered have been explained.

### Funding

Funding: This project was funded by the National Institute for Health Research (NIHR) School for Primary Care Research grant (project reference 563). The Primary Care Research Centre, University of Southampton is a member of the NIHR School for Primary Care Research and supported by NIHR Research funds. Service support costs will be paid by the CRN. CDM is funded by the National Institute for Health Research (NIHR) Collaborations for Leadership in Applied Health Research and Care West Midlands and the NIHR School for Primary Care Research. The EMPathicO e-learning package was developed using LifeGuide software, which was partly funded by the National Institute for Health Research Southampton Biomedical Research Centre (BRC). NIHR Local Clinical Research Networks (CRNs) supported practice recruitment. The views expressed are those of the authors and not necessarily those of the NIHR or the Department of Health and Social Care. The study sponsor (University of Southampton) and funders had no role in study design; collection, management, analysis, and interpretation of data; writing of the report; or the decision to submit the report for publication. The researchers are independent from the funders and all authors had full access to all of the data (including statistical reports and tables) in the study and can take responsibility for the integrity of the data and the accuracy of the data analysis.

## Supporting information

Tidier Statement

## Data Availability

All data produced in the present study are available upon reasonable request to the authors

## Acknowledgements

The TIP trial team gratefully acknowledge the contribution of our Public Advisory Group (Jennifer Bostock, Clara Martins de Barros, Mark Lamond, Manoj Mistry, Hazel Patel, and David Truswell), our Independent Trial Steering Committee (Joanne Reeve, Ian Dickerson, Ines Rombach, Philip Pallman), and our administrator (Tanya Palmer).

## Contributor and Guarantor Information

All authors meet the ICMJE criteria for authorship, i.e., have made substantial contributions to the conception or design of the work; or the acquisition, analysis, or interpretation of data for the work; AND drafted the work or reviewed it critically for important intellectual content; AND approved the version to be published; AND agree to be accountable for all aspects of the work in ensuring that questions related to the accuracy or integrity of any part of the work are appropriately investigated and resolved.

Specific contributions as per the CRediT taxonomy are: Conceptualization (FLB, TB, KG, NC, RDH, ET, AH, MER, MJR, CM, LC, JB, BS, LM, SP, JV, HA, JH, GML, JN, NI, PHL, PL, HAE); Data curation (ET); Formal analysis (ET, FLB, RDH, GML, SP); Funding acquisition (FLB, KG, MJR, CM, LC, JB, BS, LM, JV, HA, JH, GML, PL, HAE), Investigation (NC, RDH, ET, AH, MER, SP, NI), Methodology (FLB, ET, HAE), Project administration (FLB, NC, HAE, JN), Software (SP), Supervision (FLB, MJR, CM, LC, JN, HAE), Validation (BS), Writing – original draft (ET), Writing – review & editing (FLB, TB, KG, NC, RDH, AH, MER, MJR, CM, LC, JB, BS, LM, SP, JV, HA, JH, GML, JN, NI, PHL, PL, HAE).

The guarantor (F L Bishop) accepts full responsibility for the work and/or the conduct of the study, had access to the data, and controlled the decision to publish. The corresponding author attests that all listed authors meet authorship criteria and that no others meeting the criteria have been omitted.

## Copyright/License for Publication

The Corresponding Author has the right to grant on behalf of all authors and does grant on behalf of all authors, a worldwide licence to the Publishers and its licensees in perpetuity, in all forms, formats and media (whether known now or created in the future), to i) publish, reproduce, distribute, display and store the Contribution, ii) translate the Contribution into other languages, create adaptations, reprints, include within collections and create summaries, extracts and/or, abstracts of the Contribution, iii) create any other derivative work(s) based on the Contribution, iv) to exploit all subsidiary rights in the Contribution, v) the inclusion of electronic links from the Contribution to third party material where-ever it may be located; and, vi) licence any third party to do any or all of the above.

## Declaration of Competing Interest

The authors declare the following competing interests: FLB (research grant from NIHR School for Primary Care Research paid to institution; speakers honoraria from Stoneygate Centre for Empathic Healthcare and New Scientist), TB (research grant from NIHR School for Primary Care Research paid to institution), KG (research grant from NIHR School for Primary Care Research paid to institution; ModRUM license holder), NC (research grant from NIHR School for Primary Care Research paid to institution), RDH (none declared), ET (research grant from NIHR School for Primary Care Research paid to institution), AH (research grant from NIHR School for Primary Care Research paid to institution), MER (research grant from NIHR School for Primary Care Research paid to institution), MJR (research grant from NIHR School for Primary Care Research paid to institution; other research funding from NIHR SPCR, HTA, and PGfAR paid to institution; NIHR Research Professorship; on TSC/DMC for ERICA, BabyBathe and ASYMPTOMATIC trials), CM (research grant from NIHR School for Primary Care Research paid to institution; other funding from NIHR, MRC, NHS paid to institution; is the Director of the NIHR SPCR), LC (research grant from NIHR School for Primary Care Research paid to institution; paid role as Chief Medical Officer, NHS Shropshire, Telford and Wrekin; Salaried GP at Brook Medical Centre, Stoke-on-Trent; Senior Lecturer in General Practice Research, Keele University), JB (research grant from NIHR School for Primary Care Research), BS (research grant from NIHR School for Primary Care Research paid to institution; other research funding from NIHR paid to institution; member of NIHR HTA Commissioning Panel – 15/09/2020 to present), LM (research grant from NIHR School for Primary Care Research paid to institution), SP (research grant from NIHR School for Primary Care Research paid to institution), JV (none declared), HA (research grant from NIHR School for Primary Care Research paid to institution; other research funding from NIHR, Research Council of Norway, University of Warwick/eConsult Ltd paid to institution; honoraria from Imperial College London, UCL, North West Cancer Research; received support for travel from NIHR, RCGP, University of Birmingham; advisory board member/chair for NIHR159467, NIHR160384, BRACE rapid evaluation centre, HED-LINE study, EPaCCS study; is Vice Chair of the Scientific Foundation Board, Royal College of General Practitioners), JH (research grant from NIHR School for Primary Care Research paid to institution), GML (none declared), JN (none declared), NI (none declared), PHL (none declared), PL (research grant from NIHR School for Primary Care Research paid to institution), HAE (research grant from NIHR School for Primary Care Research paid to institution; is Deputy Academic Capacity Development Lead for the NIHR SPCR and sits on the NIHR SPCR Board and Exec; works clinically as a GP at New Horizons Medical Partnership in Southampton as part of Professor of Primary Care Research post at the University of Southampton; Co-Authors the Oxford Handbook of General Practice published by Oxford University Press).

## Funding

NIHR School for Primary Care Research grant 563.

## Notes

### Author Declarations

South Central - Hampshire B Research Ethics Committee gave ethical approval for this work (ref: 22/SC/0145)

